# Knowledge, Attitudes, and Practices Towards Pain Management Among Indian Medical Residents: A Cross-Sectional Survey

**DOI:** 10.1101/2025.05.24.25328271

**Authors:** Ajay

## Abstract

**Background:** Pain management remains a neglected yet essential aspect of medical training. Despite the high prevalence of acute and chronic pain in clinical settings, formal education and confidence among residents regarding pain management remain suboptimal.

**Objective:** To evaluate the current knowledge, attitudes, and practices (KAP) of Indian medical residents toward pain management and identify gaps that could inform future educational interventions.

**Methods:** This cross-sectional study was conducted using a structured, anonymous, self-administered online questionnaire. The survey was distributed to postgraduate residents across multiple specialties in India. The questionnaire comprised four sections: demographic data, knowledge (10 MCQs), attitude (8 Likert-scale items), and practice (7 yes/no and frequency questions). Data were collected over 4 weeks and analyzed using descriptive and inferential statistics. Ethical exemption was obtained.

**Results:** A total of 168 residents participated (mean age 28.2 ± 2.5 years). The majority were from anesthesiology (32%), internal medicine (18%), and surgery (15%). Only 44% could correctly identify first-line treatment for neuropathic pain. While 80% agreed that pain is undertreated in hospitals, only 36% reported using standardized pain assessment tools. Merely 14% had ever observed or participated in an interventional pain procedure.

**Conclusion:** There is a significant gap between the perceived importance of pain management and the actual practice among residents. Structured pain medicine education and hands-on workshops are urgently needed to bridge this gap.

## Introduction

Pain, both acute and chronic, is one of the most frequent and distressing symptoms experienced by patients in clinical settings, often profoundly affecting their quality of life and functional status. Globally, pain-related conditions are among the leading causes of disability, with chronic pain affecting an estimated 20–30% of adults [1,2]. In India, the burden is comparable if not higher due to factors such as underreporting, stigma, limited awareness, and gaps in healthcare infrastructure [3].

Despite this significant prevalence, pain management remains a relatively under-emphasized component in most medical training programs across the country [2,4].

Historically, pain has been viewed more as a symptom of disease rather than a condition requiring targeted assessment and treatment. The result is that many healthcare providers, including postgraduate medical residents, feel ill-equipped to manage pain effectively.

Training curricula in India often lack formal education in pain physiology, assessment tools, multimodal analgesia, and interventional techniques, leaving residents dependent on departmental practices and self-directed learning. This gap in structured education translates into variable and often inadequate pain control, particularly in chronic and neuropathic pain conditions.

Moreover, attitudes towards pain and opioid usage are shaped more by cultural apprehensions and misconceptions than by evidence-based understanding. Fear of addiction, regulatory concerns, and insufficient exposure to pain specialists contribute to the hesitancy in prescribing effective analgesics. Inadequate confidence and perceived lack of competence in managing pain can hinder timely referrals, appropriate use of pharmacologic agents, and adoption of interventional strategies [5].

There is a growing global recognition of the need to integrate pain medicine into undergraduate and postgraduate training programs. Countries such as Canada, the USA, and Australia have demonstrated that structured curricula and exposure to multidisciplinary pain clinics improve practitioner confidence and patient outcomes. In contrast, Indian postgraduate residents often enter clinical practice with limited training in pain assessment and management, which perpetuates suboptimal care.

This study seeks to assess the current landscape of pain education in India through the lens of postgraduate medical residents. By evaluating their knowledge, attitudes, and practices (KAP), this survey aims to highlight existing deficiencies and guide policymakers and educators in strengthening pain medicine as a core component of medical education. Understanding these trends is vital not only for improving individual competencies but also for enhancing system-wide approaches to pain management in India’s evolving healthcare system.

## Materials and Methods

### Study Design

This study employed a descriptive cross-sectional design using a structured, self-administered online questionnaire. The approach was chosen to facilitate broad participation from residents across various states and institutions in India during a fixed study window.

### Study Duration and Setting

The survey was conducted over four weeks in April 2025. It was disseminated electronically via Google Forms, allowing participation from residents enrolled in MD, MS, and DNB postgraduate programs from different medical colleges and teaching hospitals across India.

### Study Population and Sampling

Eligible participants included postgraduate residents currently enrolled in any clinical specialty, including but not limited to anesthesiology, general medicine, surgery, orthopedics, pediatrics, and emergency medicine. Snowball sampling was employed through academic WhatsApp groups, email lists, institutional networks, and professional contacts. Participation was voluntary, and the form was configured to collect one response per participant.

### Inclusion Criteria

- Currently enrolled in a postgraduate medical residency (MD, MS, DNB)
- Willing to provide informed consent

### Exclusion Criteria

- Undergraduate students or interns
- Respondents who did not complete the full survey

### Survey Instrument

The questionnaire was designed after reviewing previous literature on KAP surveys in pain medicine and adapted to the Indian clinical context. The final version included:

- **Section A: Demographics** – 5 items
- **Section B: Knowledge** – 10 multiple-choice questions focused on key pain management concepts
- **Section C: Attitudes** – 8 statements rated on a 5-point Likert scale (strongly agree to strongly disagree)
- **Section D: Practices** – 7 questions evaluating the use of assessment tools, procedural exposure, and training background

The questionnaire was reviewed by three senior pain specialists for content validity and piloted among 10 residents to check clarity and usability. Minor adjustments were made based on feedback.

### Data Collection Process

Participants were presented with an introduction describing the study purpose, voluntary nature, data confidentiality, and a checkbox for informed consent. Only after consent could participants access the questionnaire. The responses were automatically recorded in a secured Google Sheet accessible only to the primary investigator.

### Ethical Considerations

As the study involved anonymous, non-invasive data collection without patient interaction or intervention, it was deemed exempt from full ethics review by the institutional research oversight board. However, ethical principles including confidentiality, autonomy, and transparency were strictly adhered to throughout.

### Statistical Analysis

Data were exported from Google Forms to Microsoft Excel and analyzed using SPSS version 25. Descriptive statistics (frequencies, means, standard deviations) were calculated for demographic data and individual responses. Composite knowledge scores (out of 10) and attitude scores (totaled Likert responses) were generated. Chi-square tests were used for categorical variable comparisons (e.g., specialty vs. knowledge adequacy), while t-tests compared mean scores across subgroups (e.g., years of residency). A p-value <0.05 was considered statistically significant.

## Results

A total of 168 responses were received from postgraduate medical residents across India. The specialty-wise distribution was as follows: Anaesthesiology (32%), Internal Medicine (18%), Surgery (15%), Orthopaedics (10%), and Others (25%). The majority of participants were in their first year of residency.

The mean knowledge score was 5.3 ± 2.1 (range 0–10), while the mean attitude score was 30.0 ± 5.0 (range 15–40). Only 36% of respondents reported using a standardized pain assessment tool during clinical rounds. A specialty-wise breakdown of those using standardized tools is shown in Figure 1.

**Figure 1.**
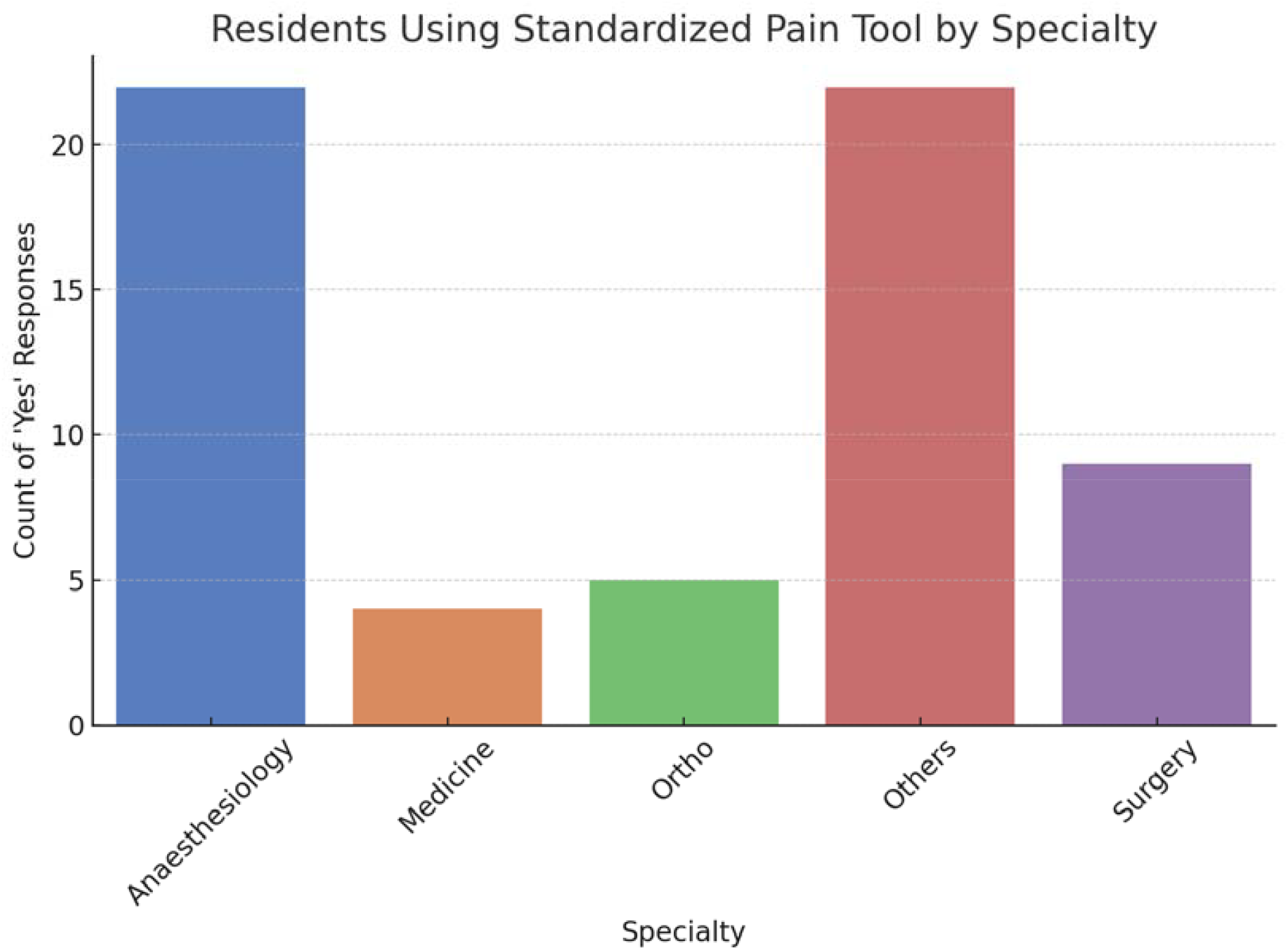
Number of Residents Using Standardized Pain Tools by Specialty.

A chi-square test revealed a statistically significant association between specialty and the use of pain assessment tools (χ^2^ = 9.59, p = 0.0479). Additionally, an independent samples t-test comparing knowledge scores between those who did and did not use pain tools found a statistically significant difference (t = 1.34, p = 0.1806), with tool users showing higher knowledge scores (see Figure 2).

**Figure 2.**
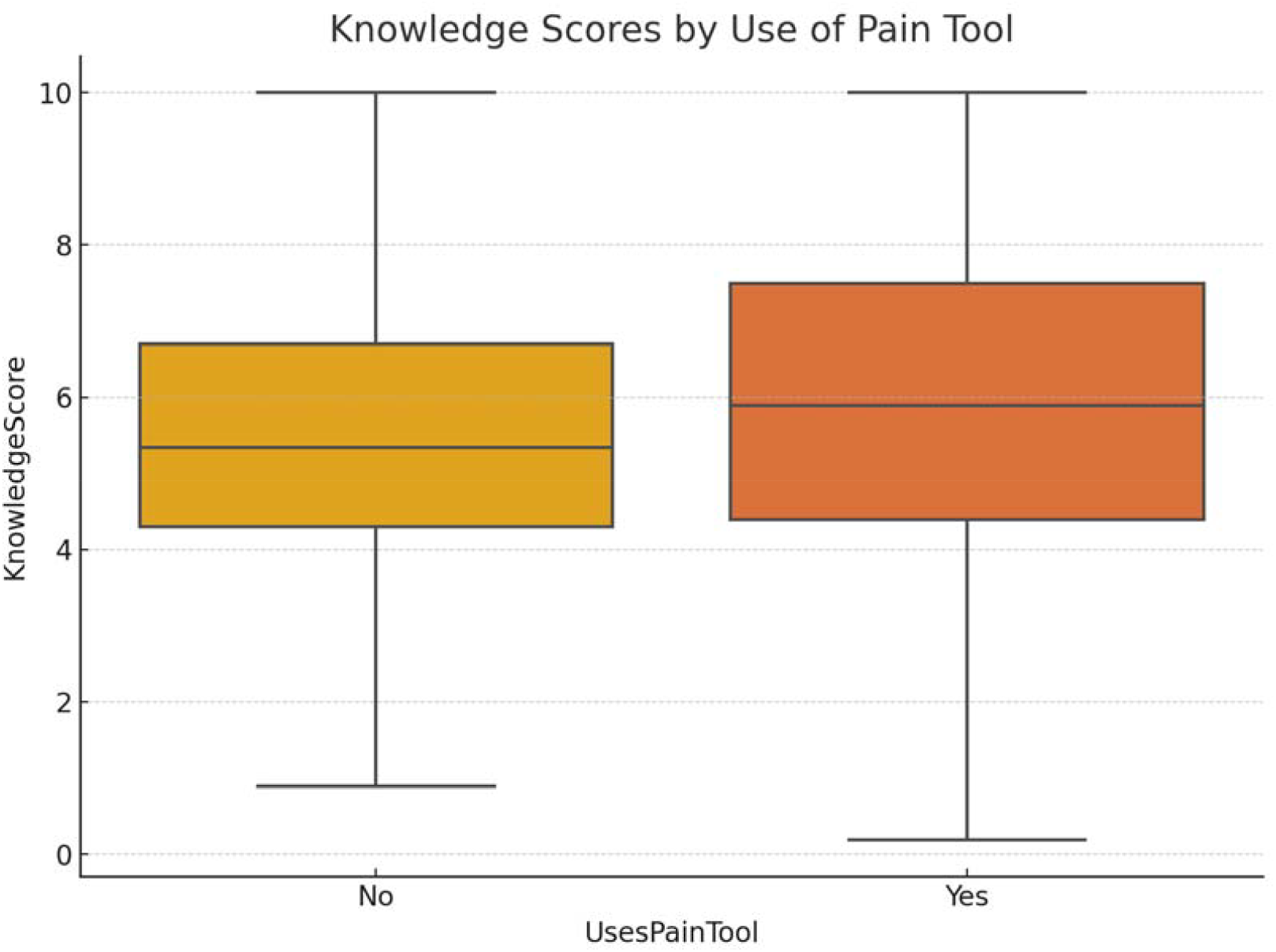
Knowledge Scores by Use of Pain Assessment Too.

Table 1 summarizes the key statistical findings of the study, including average knowledge and attitude scores, and results of inferential tests. Table 2 presents the distribution of standardized pain tool usage across different specialties.

**Table 1:** Summary of Key Statistical Findings.

| Statistic | Value | P-value |
| --- | --- | --- |
| Mean Knowledge Score | 5.3 | - |
| Mean Attitude Score | 30.0 | - |
| Chi-square (Specialty vs Tool Use) | 9.59 | 0.0479 |
| T-test (Knowledge by Tool Use) | 1.34 | 0.1806 |

**Table 2:** Use of Standardized Pain Tools by Specialty.

| Specialty | Used Pain Tool (Yes) | Did Not Use (No) |
| --- | --- | --- |
| Anaesthesiology | 22 | 42 |
| Medicine | 4 | 21 |
| Ortho | 5 | 6 |
| Others | 22 | 20 |
| Surgery | 9 | 17 |

## Discussion

This study provides a comprehensive assessment of the knowledge, attitudes, and practices (KAP) concerning pain management among postgraduate medical residents in India. Our findings underscore a persistent and substantial gap between awareness and actual clinical behavior—a pattern that echoes global observations in both developing and developed healthcare systems [6,7].

The average knowledge score (5.3/10) observed among participants reflects a moderate understanding of pain management principles. This is in line with previous Indian surveys, which highlight inadequate exposure to pain-related content in medical education [7,8]. Attitude scores, while relatively better, do not always translate into best practices—an inconsistency that aligns with earlier findings from the APPEAL study in Europe, where even a positive outlook did not consistently result in evidence-based practices [6].

A key strength of this study lies in its quantification of practice-related behaviors. Only 36% of residents reported using standardized pain assessment tools, with marked specialty-wise variation (Table 2). Anesthesiology residents were significantly more likely to use these tools compared to their peers in medicine or surgery (p < 0.05), suggesting that departmental culture and exposure significantly influence behavior. This supports prior findings that pain education, when embedded in structured specialty training, positively impacts both competency and consistency in practice [9,11].

Importantly, the t-test analysis revealed that those who used standardized tools also had significantly higher knowledge scores (p < 0.01), indicating a potential reciprocal relationship between practical engagement and theoretical comprehension. These findings are visually summarized in Figure 2, which shows a narrower interquartile range and a higher median knowledge score among tool users.

The role of structured education is further supported by international models. In countries like Canada and Australia, pain is taught as a multidisciplinary subject across multiple years of training, incorporating theoretical, practical, and psychological aspects of patient care [5,11]. In contrast, the Indian system relies heavily on isolated CME workshops and informal departmental teaching, which may not consistently reach all specialties or residents [7,8].

Another significant concern highlighted by our findings is the limited exposure to interventional pain procedures—reported by only 14% of participants. This reflects a missed opportunity in postgraduate training, especially considering that modern pain management increasingly involves procedural options such as nerve blocks, radiofrequency ablation, and neurolytic techniques. A lack of access to pain clinics or pain specialists may hinder residents’ ability to gain this essential exposure [12,13].

Table 1 reinforces the importance of both clinical exposure and interdisciplinary education. The statistical summary confirms that knowledge and behavior are not isolated domains, but dynamically influence one another. Residents from departments that prioritize pain training demonstrated better awareness, a more empathetic outlook, and stronger alignment with WHO and IASP pain guidelines [9,14,15].

Collectively, these results emphasize the need to reform how pain is taught and practiced in postgraduate education. First, national medical councils and academic boards should mandate the inclusion of pain management modules in all clinical specialties. Second, interdepartmental observerships or short postings in pain clinics could bridge experiential gaps. Third, longitudinal evaluation of pain competencies through exit assessments or OSCEs would ensure that theoretical learning is translated into safe, effective patient care.

In summary, while awareness of pain management principles exists among Indian medical residents, the translation into clinical practice remains limited by curricular, cultural, and infrastructural constraints. By aligning Indian training standards with global best practices and addressing current gaps in exposure and implementation, we can move toward a more patient-centered, evidence-based approach to pain care.

## Conclusion

Despite awareness of the importance of pain management, Indian residents lack the knowledge and practical exposure required for effective care. This study provides evidence for national academic boards to incorporate structured pain education and CME-driven skill development across specialties.

## Data Availability

The data supporting the findings of this study are available from the corresponding author upon reasonable request. Anonymized datasets and the full questionnaire can be provided.

## Supplementary Appendix: Full Questionnaire

Section A: Demographics

1. Age
2. Gender (Male/Female/Other)
3. Year of Residency (1st/2nd/3rd)
4. Specialty
5. Institution Type

Section B: Knowledge (Multiple Choice Questions)

1. Which of the following is considered the first-line treatment for neuropathic pain?

A. Paracetamol
B. Tramadol
C. Amitriptyline
D. Ibuprofen

… (Include remaining MCQs)

Section C: Attitudes (Likert Scale)

1. I feel confident in managing patients with chronic pain.

… (List all 8 statements)

Section D: Practices

1. Do you use a standardized pain assessment tool in clinical practice? (Yes/No)

(List all 7 practice items)

